# Country-Wide Agent-Based Epidemiological Modeling Using 17 Million Individual-Level Microdata

**DOI:** 10.1101/2024.05.27.24307982

**Authors:** Ahmad Hesam, Frank P Pijpers, Lukas Breitwieser, Peter Hofstee, Zaid Al-Ars

## Abstract

Calibration is a crucial step in developing agent-based models. Agent-based models are notorious for being difficult to calibrate as they can express various degrees of freedom when model parameters are unknown. Models that appear correctly calibrated to match macro-level observed data perform poorly when micro-level insights need to be inferred. As a result, policymakers cannot be certain that an agent-based model can accurately describe the dynamics of the real-world phenomena that the model tries to mimic. To begin tackling this challenge, we developed a methodology for an epidemiological use case at a full population scale of 17 million agents to observe the effects of using microlevel data for the calibration on the accuracy of the microlevel model outcomes. We show that by calibrating a model on national statistics, but using individual-level microdata, we can on average get 36% more accurate model outcomes on a subnational level. Our model implementation performs two orders of magnitude faster than prior work and allows efficient calibration on HPC computer systems.

## Introduction

Agent-based modeling (ABM) has emerged as a powerful tool for exploring the behavior of complex social systems over time ***Conte and Paolucci (2014***). Agent-based models are designed to simulate the interactions between autonomous agents, allowing for a nuanced representation of emergent phenomena. In contrast to equation-based approaches, ABM allows us to program each agent to follow a set of rules that follow from a real-world system. Agents typically interact with a limited number of neighboring agents. In social systems, for example, agents are often connected through social networks (e.g. family, co-workers, friends).

Practical application of ABM to real-world use cases faces several challenges. One of the main difficulties in ABM is dealing with the extensive parameter space arising from the diverse free parameters attributed to each agent and their behaviors ***Crooks et al. (2008***). These parameters encompass a wide array of individual attributes and interactions, adding substantial complexity to the model. Moreover, obtaining datasets that provide precise, detailed information at the individual agent level (i.e. *microdata*) can be rather challenging. Even in cases where such a dataset is available, limitations in computational capacity often impede the simulation of each real-world actor as an individual agent in the model. The sheer amount of computational resources required to manage a large number of agents can be prohibitive, making it difficult to achieve the desired level of precision and fidelity in the model’s representation of real-world dynamics. As a result, many ABM studies let one simulated agent represent multiple real-world actors (e.g. one agent represents 100 persons) ***Edmonds and Meyer (2017***). Throughout this paper we will refer to the ratio between simulated agents and real-world actors is often referred to as the *ontological correspondence*. We found no studies that discuss and experimentally explore the effects of varying the ontological correspondence in agent-based modeling.

In this work, we analyze the effects of varying the ontological correspondence of ABMs on the simulated results, specifically highlighting the impact when simulated agents represent individual real-world actors. First, we investigate how high-quality social microdata affects the output results of agent-based simulation. We also show how the use of a state-of-the-art agent-based simulation platform (that can harness the computational power of modern hardware) enables us to scale up to high-resolution ABMs, as well as identify values for the diverse free parameters attributed to each agent. The platform BioDynaMo ***Breitwieser et al. (2021***) focuses on supporting high-performance and modular agent-based simulations. BioDynaMo demonstrated unprecedented performance in biomedical applications, efficiently executing simulations of up to 1 billion agents on a single server ***Breitwieser et al. (2023***).

The relevant social microdata for epidemiological modeling, (e.g. age, sex, relationships, employment status, etc.) are often sensitive data. The use of such data is understandably subject to rigorous regulations such as the EU GDPR (General Data Protection Regulation). For this reason in this paper, particular attention and extra emphasis is given to the secure supercomputing method we use to access such data in this study.

In this work we present the following contributions:

- We implement a country-wide epidemiological model for the Netherlands in BioDynaMo on a 1:1 agent-to-person resolution. At the time of the first wave of the COVID-19 epidemic in early 2020, the Netherlands had about 17.4 million inhabitants.
- We construct a method to simulate the model on a secure supercomputer platform to perform parameter space exploration using high-quality sensitive microdata using a total of 1536 CPU cores.
- We present an analysis of the effects of using various agent-to-person model resolutions and real-life microlevel data versus randomized synthetic data. We also present the results of a parameter sensitivity analysis for various resolutions.
- We describe a methodology behind distributing the calibration of agent-based models using the particle swarm optimization algorithm as part of a high-performance agent-based modeling framework.

## Results

### Hospital admission resolution effects

A key result of our study is the comparison between the observed hospital admissions during the initial COVID-19 wave in the Netherlands and its simulated counterpart. This comparison, detailed across various agent-to-person ratios, is shown in Figure 1. Across all examined ratios, there is a notable congruence between the simulated hospital admissions and the actual observed data, indicating the model’s robustness in mirroring real-world outcomes. To quantify the accuracy improvement, we calculate the percentage difference in the root mean squared error between the observed data and the mean simulated data for each resolution. We find an accuracy improvement of 49.3% when increasing the resolution from 1:100 to 1:10, and an improvement of 1.7% when increasing the resolution from 1:10 to 1:1.

**Figure 1.**
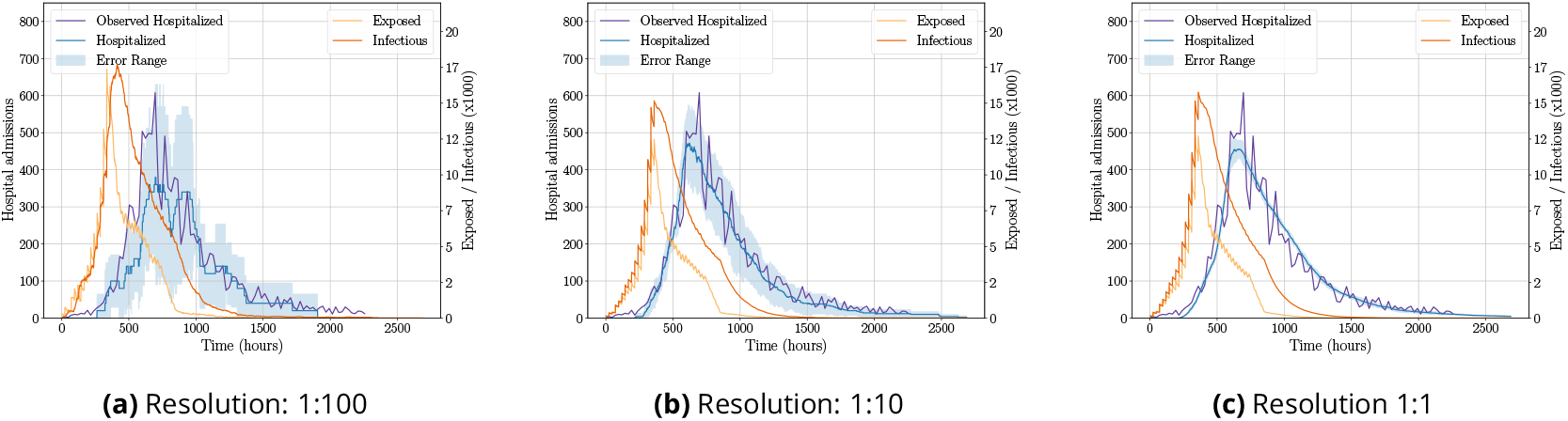
National hospital admissions in the Netherlands during the first wave of COVID-19 in 2020. Left y-axis: the daily hospital admissions from simulation (dark purple) and observed data (light purple). Right y-xis: the daily number of infections and exposed people (simulated data) in orange and yellow, respectively.

However, it is important to highlight that the simulated results for the 1:100 agent-to-person ratio exhibit a relatively wide error margin. As we refine the agent-to-person ratio, leading to higher resolution simulations, we witness a strong reduction in this error margin, suggesting an improvement in the model’s precision. Interestingly, the 1:100 ratio simulations depicted in Figure 1a reveal a potential double-peak phenomenon, which diverges from the actual single-peak pattern observed in reality. This discrepancy underscores the limitations of certain resolutions in accurately capturing the pandemic’s dynamics. Additionally, for the metrics related to exposed and infectious categories, our model was able to generate outcomes in the absence of real-world comparative data.

### Local hospital admission accuracy

Another key finding of our study is exposing the influence of microdata on the accuracy of hospital admissions on a local (municipality) level during the initial wave of COVID-19 for a selected set of municipalities in the Netherlands: Eindhoven, Groningen, and The Hague. Figure 2 displays the outcomes derived from calibrating and simulating our model with synthetic population data, while Figure 3 displays the outcomes using individual-level registry data. This comparison is designed to show the effects of using detailed individual data on a local scale. We observe a notable difference in the accuracy of the results; the use of microdata yields outcomes that align more closely with the actual hospital admissions compared to the simulations based on synthetic data. To quantify the accuracy improvement, we calculate the percentage difference in the root mean squared error between the observed data and the mean simulated data for each municipality. We find that applying microdata improves model accuracy by 36.2% for Eindhoven, 22.9% for Groningen, and 48.0% for The Hague, resulting in a mean accuracy improvement of 35.7%. Additionally, the variability of the results, as indicated by the error range, is significantly reduced when microdata is utilized. These findings highlight the potential benefits of incorporating detailed individual-level data in agent-based modeling studies.

**Figure 2.**
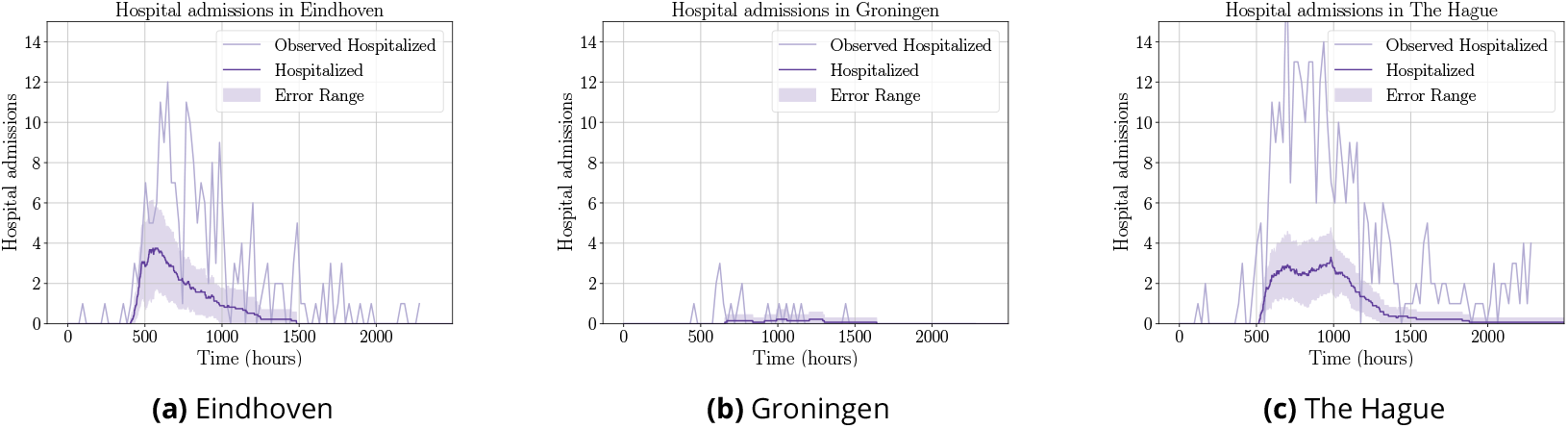
Local hospital admissions in the municipalities of (a) Eindhoven, (b) Groningen, and (c) The Hague with the use of synthetic population data during calibration and simulation.

**Figure 3.**
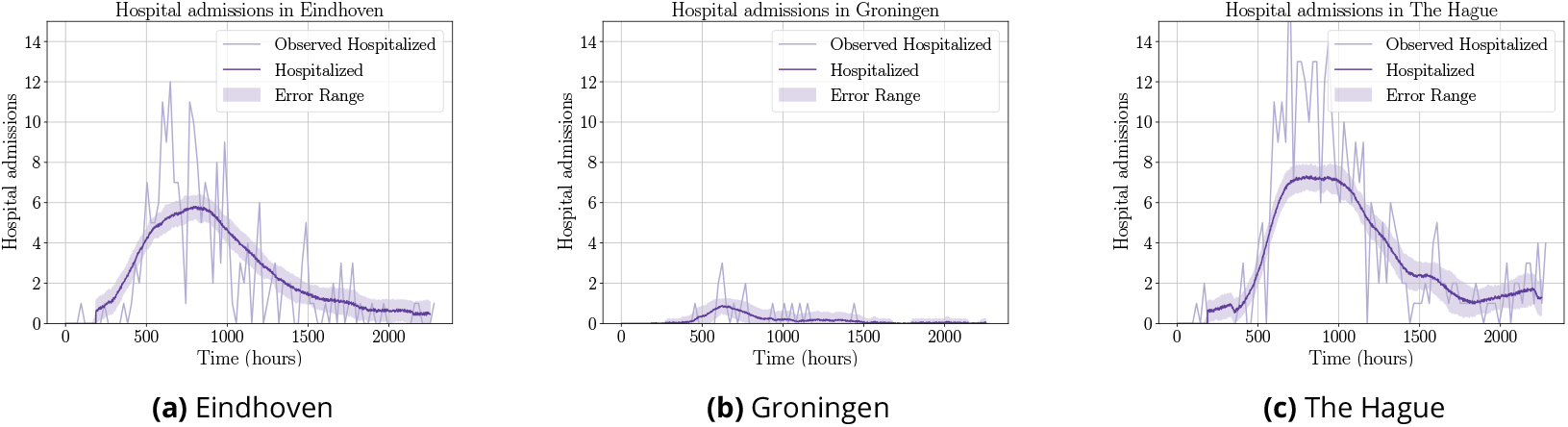
Local hospital admissions in the municipalities of(a) Eindhoven, (b) Groningen, and (c) The Hague with the use of CBS microdata population data during calibration and simulation

### Parameter sensitivity analysis

The outcomes of the parameter sensitivity analysis are detailed in Figure 4, where we subtly adjust two of the four variable parameters (beta1 and beta2) in our model to assess their impact on the stability of our results. Specifically, we examine fluctuations in two key aspects: 1) the full-width at half maximum (FWHM) and 2) the amplitude of the wave-like pattern, as depicted in Figure 1. Our analysis reveals that the results evolve gradually as the parameters change, indicating a stable and proportionate influence on the outcomes. This stability suggests that the parameters are well-defined and do not exert undue influence on the model’s predictions, thereby bolstering our confidence in the parameters’ validity and their role within the model. The effects of the other two free parameters beta3 and beta4) that were not added in this section can be found in the appendix. These two beta values represent the phase-bound behaviors such as wearing face masks and social distancing in the third and fourth phases of the first COVID-19 wave in the Netherlands.

**Figure 4.**
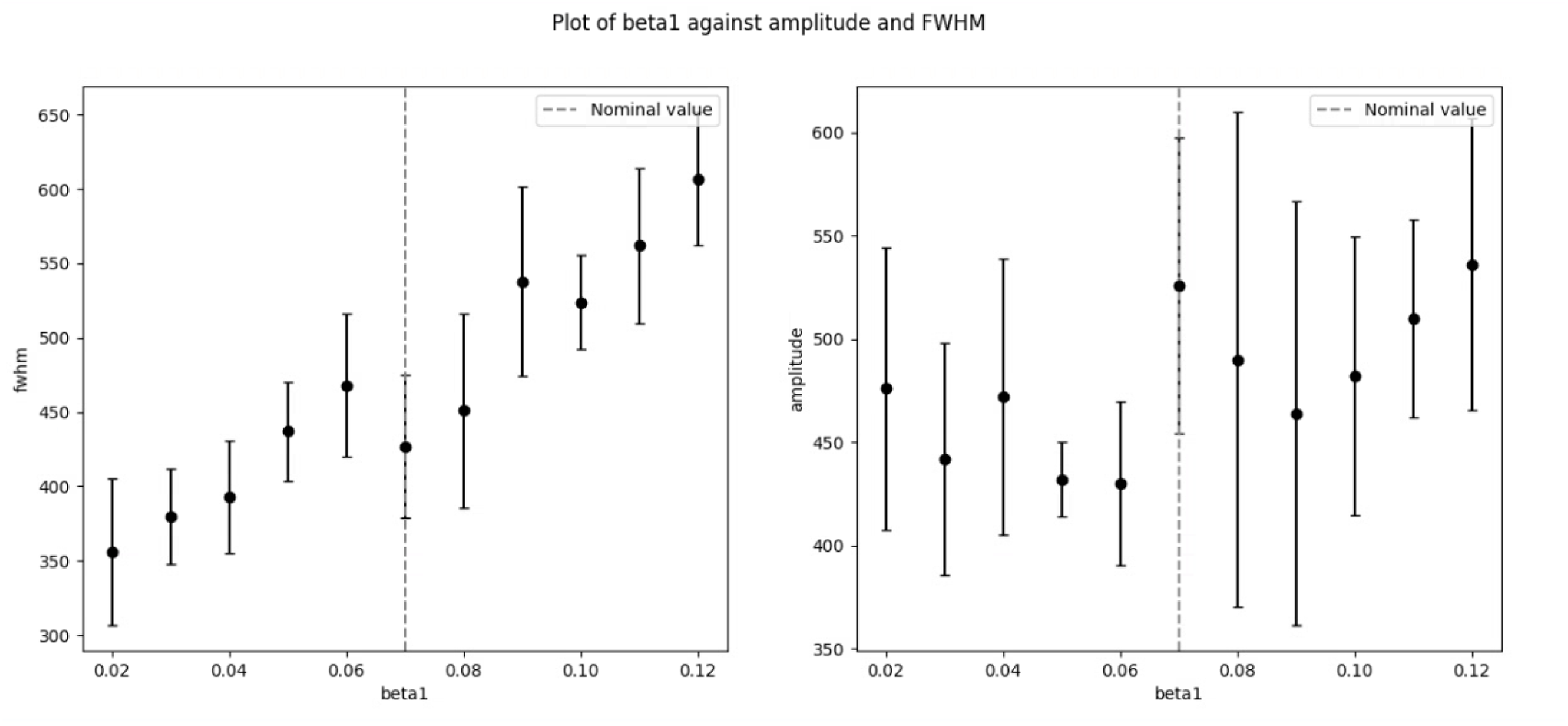
Parameter sensitivity analysis using the one-factor-at-a-time (OFAT) method for the `beta1` parameter.

**Figure 5.**
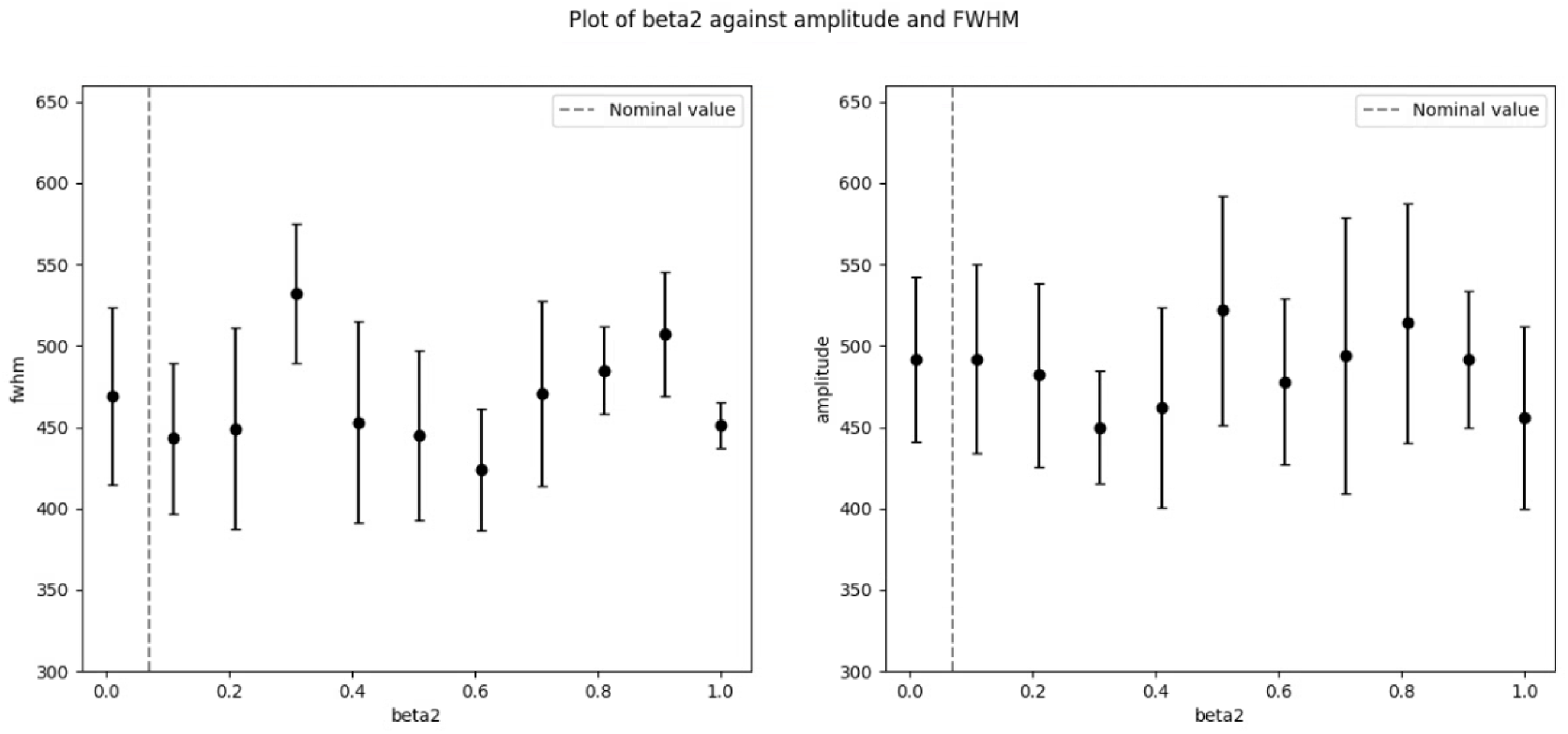
Parameter sensitivity analysis using the one-factor-at-a-time (OFAT) method for the `beta2` parameter.

### Computational performance

We wish to highlight the computational performance of our simulations, an aspect often overlooked or briefly touched upon in discussions of agent-based models. Our decision to focus on computational efficiency stems from the pivotal role the BioDynaMo framework played in enabling our research. The advanced computational capabilities of BioDynaMo ***Breitwieser et al. (2023***); ***Hesam et al. (2021***) significantly contributed to the feasibility of our study, allowing us to undertake complex simulations that would otherwise have been unattainable. The details on the hardware that was used for the performance benchmarks results can be found in Table 1. The results of the performance benchmarks can be found in Figure 6.

**Table 1.**
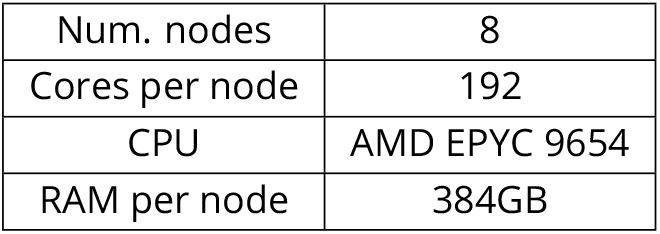
System specifications for simulations. Details the computing resources used in our simulation runs. The nodes are part of Snellius, the Dutch national supercomputer cluster.

**Figure 6.**
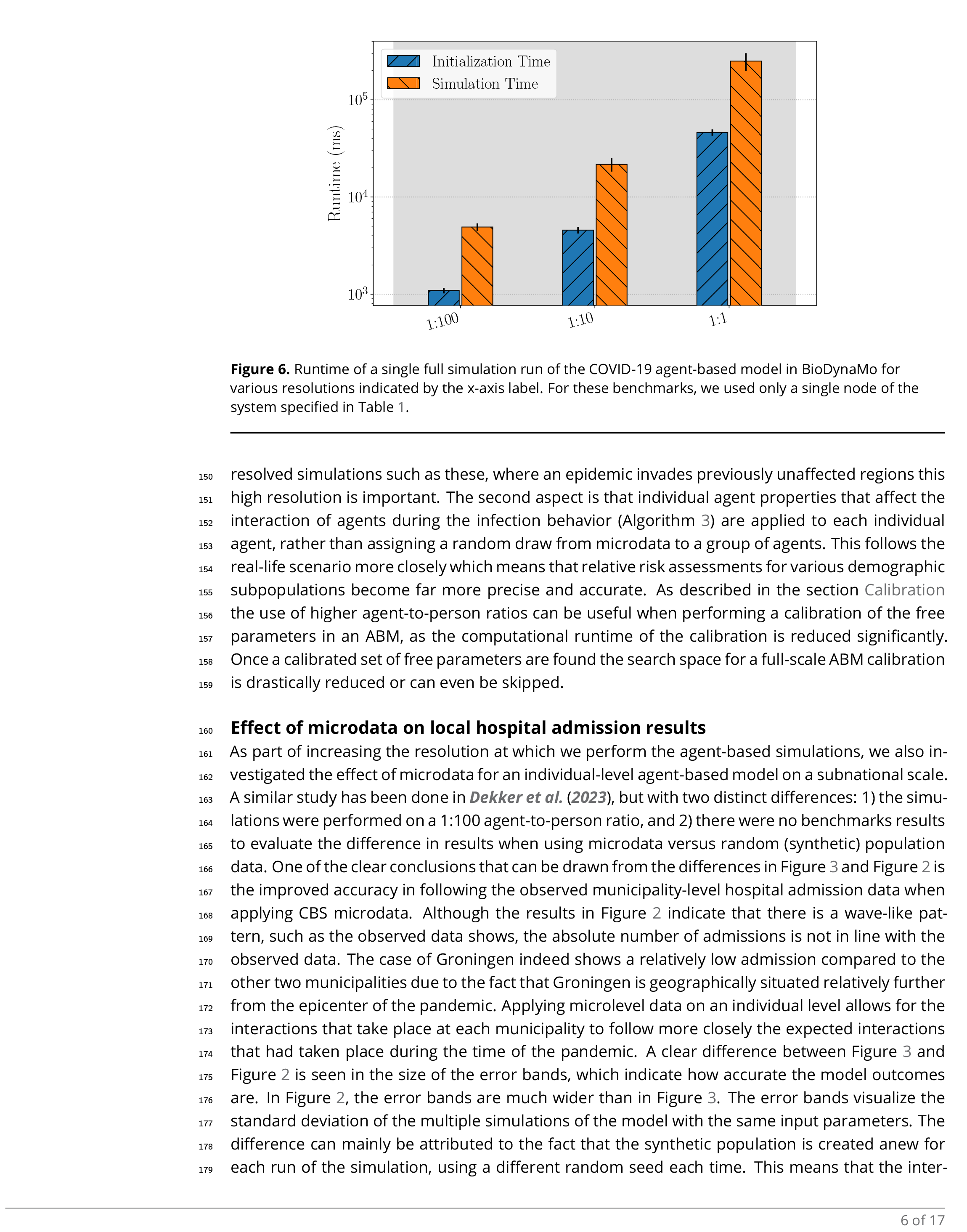
Runtime of a single full simulation run of the COVID-19 agent-based model in BioDynaMo for various resolutions indicated by the x-axis label. For these benchmarks, we used only a single node of the system specified in Table 1.

The total duration of a simulation can be divided into initialization time and simulation time. During initialization, we create agent objects, assign attributes and behaviors to the agents, and apply configuration parameters. During the simulation, we iterate over all timesteps and all agents in each timestep. Initialization time can be significant, as agent attributes need to be read from a file and are therefore limited by disk reading speed.

The work of ***Dekker et al. (2023***) reported a runtime of 20 hours for a single full simulation run on a single node where one agent represents one person. Our work demonstrates that the same model can be simulated in less than 5 minutes on a single node, achieving a speedup of 240×. Although we do not know the specific hardware used in their benchmarks, they claimed that computation time was the bottleneck for simulating their model at such a high resolution. BioDynaMo’s ability to utilize modern hardware capabilities eliminates this issue.

## Discussion

### Hospital admission resolution effect

The observation of a decrease in the width of the error bands surrounding the simulated hospital admissions in Figure 1 confirms our initial hypothesis, in which we stated that a lower agent-to-person ratio would more accurately track the consequences of the social phenomena at hand. We can attribute this observation to two aspects of our model. The first aspect is that a single agent-to-agent infection increments the infection by 1 person rather than 10 or 100, which allows for more granular stochastic behavior to occur at each simulation step. Especially in regionally-resolved simulations such as these, where an epidemic invades previously unaffected regions this high resolution is important. The second aspect is that individual agent properties that affect the interaction of agents during the infection behavior (Algorithm 3) are applied to each individual agent, rather than assigning a random draw from microdata to a group of agents. This follows the real-life scenario more closely which means that relative risk assessments for various demographic subpopulations become far more precise and accurate. As described in the section Calibration the use of higher agent-to-person ratios can be useful when performing a calibration of the free parameters in an ABM, as the computational runtime of the calibration is reduced significantly. Once a calibrated set of free parameters are found the search space for a full-scale ABM calibration is drastically reduced or can even be skipped.

### Effect of microdata on local hospital admission results

As part of increasing the resolution at which we perform the agent-based simulations, we also investigated the effect of microdata for an individual-level agent-based model on a subnational scale. A similar study has been done in ***Dekker et al. (2023***), but with two distinct differences: 1) the simulations were performed on a 1:100 agent-to-person ratio, and 2) there were no benchmarks results to evaluate the difference in results when using microdata versus random (synthetic) population data. One of the clear conclusions that can be drawn from the differences in Figure 3 and Figure 2 is the improved accuracy in following the observed municipality-level hospital admission data when applying CBS microdata. Although the results in Figure 2 indicate that there is a wave-like pattern, such as the observed data shows, the absolute number of admissions is not in line with the observed data. The case of Groningen indeed shows a relatively low admission compared to the other two municipalities due to the fact that Groningen is geographically situated relatively further from the epicenter of the pandemic. Applying microlevel data on an individual level allows for the interactions that take place at each municipality to follow more closely the expected interactions that had taken place during the time of the pandemic. A clear difference between Figure 3 and Figure 2 is seen in the size of the error bands, which indicate how accurate the model outcomes are. In Figure 2, the error bands are much wider than in Figure 3. The error bands visualize the standard deviation of the multiple simulations of the model with the same input parameters. The difference can mainly be attributed to the fact that the synthetic population is created anew for each run of the simulation, using a different random seed each time. This means that the interactions within each municipality vary more from one simulation to the next, leading to a broader spread in the results across different runs.

### Parameter sensitivity analysis

Utilizing the one-factor-at-a-time (OFAT) method for sensitivity analysis reveals that the agent-based model’s free parameters exhibit low sensitivity to minor adjustments in their values. We deliberately varied the input parameters subtly to observe their effects on the model outcomes, rather than to explore the relationship between the parameters and the outcomes. In the context of the epidemiological use case applied throughout this study, the four free parameters represent the four phases of interventions during the COVID-19 pandemic in the Netherlands. Each phase corresponds to a specific set of restrictions imposed on the Dutch population during the initial wave of the COVID-19 pandemic. As shown in Results the two parameters representing the first two phases turned out to have the most influence on the model’s outcome. The first two phases can be seen as the most critical and impactful phases as they correspond to the initial outbreak and the rapid escalation of infection rates. These early stages required swift and stringent interventions to curb the virus’s spread and prevent overwhelming the healthcare system.

### General applicability

The work presented in this paper focuses on the recent COVID-19 pandemic, utilizing and refining the model from ***Dekker et al. (2023***). We demonstrated that by applying microdata at an individual level resolution, our model could achieve a finer accuracy in outcomes than the granularity of the data used for its calibration. The applicability of this finding to other models in different fields hinges on several factors. Firstly, the efficacy of the agent-based model is critically linked to the precision of the agent attribute data. In our analysis, the significant factor was the dependency of agent interactions on the demographic characteristics of each municipality. We anticipate that, in fields like urban studies, agent-based models would similarly benefit from microdata, which allows for a more accurate representation of urban populations and their dynamics, such as migration patterns and clustering phenomena. Secondly, it is essential that agents within the model can be mapped on a one-to-one basis with their real-world counterparts. This fidelity ensures that each model agent represents an individual entity in the real world, and enables the application of microdata to enhance the model. To solidify the general applicability of our findings, a follow-up study is required.

## Methods and Materials

In this section, we describe the use case model and how we calibrated the free parameters on the available data from Dutch health institutes. We perform a parameter sensitivity analysis on the found parameter set values. Furthermore, we describe the computational approach used to simulate the large-scale models.

### Datasets

The datasets used in this work are described in the Table 2.

**Table 2.** Overview of datasets used in this study. Throughout this article, we refer to a dataset by the name in this table.

| Name | Description | Reference |
| --- | --- | --- |
| POLYMOD | Age-stratified mixing matrices used to infer contact matrices per municipality and demographic group | *Prem et al. (2017)* |
| PIENTER | Quantifies the impact of physical distancing measures against COVID-19 on contacts and mixing patterns in the Netherlands. | *Backer et al. (2021)* |
| Google Mobility | Quantifies the impact of COVID-19 restrictions on the mobility of Dutch citizens | *Google (2020)* |
| CBS Microdata | National Dutch register data of 2020 containing the attributes sex, age, work status, education, and residence municipality | Statistics Netherlands<sup>1</sup> |
| NICE | Hospital admission data per municipality per day in the Netherlands | *Nationale Intensive Care Evaluatie (NICE) (2020)* |

### Synthetic population data

In our study, as an alternative to the CBS microdata we generate synthetic population data for two primary purposes: firstly, to prototype the model, and secondly, to serve as a baseline for evaluating the impact of microdata integration. The confidential nature of the CBS microdata necessitates conducting all related processing within a secure computing environment, as detailed in Secure Supercomputing Infrastructure. To facilitate the functional development of the agent-based model outside this restricted setting, we need to work with the synthetic population data. This approach proved particularly advantageous for exploratory, trial-and-error methodology, enabling us to efficiently identify the necessary tools and software packages for constructing the final model. We use the same synthetic population data to perform the comparative study between synthetic data and CBS microdata and their contrasting effects on the model outcomes.

The pseudocode on how we generate the synthetic population can be found in Algorithm 1. In short, we determine the number of individuals per demographic group from the publicly known statistics from Statline ***Centraal Bureau voor de Statistiek (2023***), a public tool from Statistics Netherlands (CBS). For each individual of the group we assign random values for the sex, age, and home location (i.e. municipality).

#### Algorithm 1

Create synthetic population data

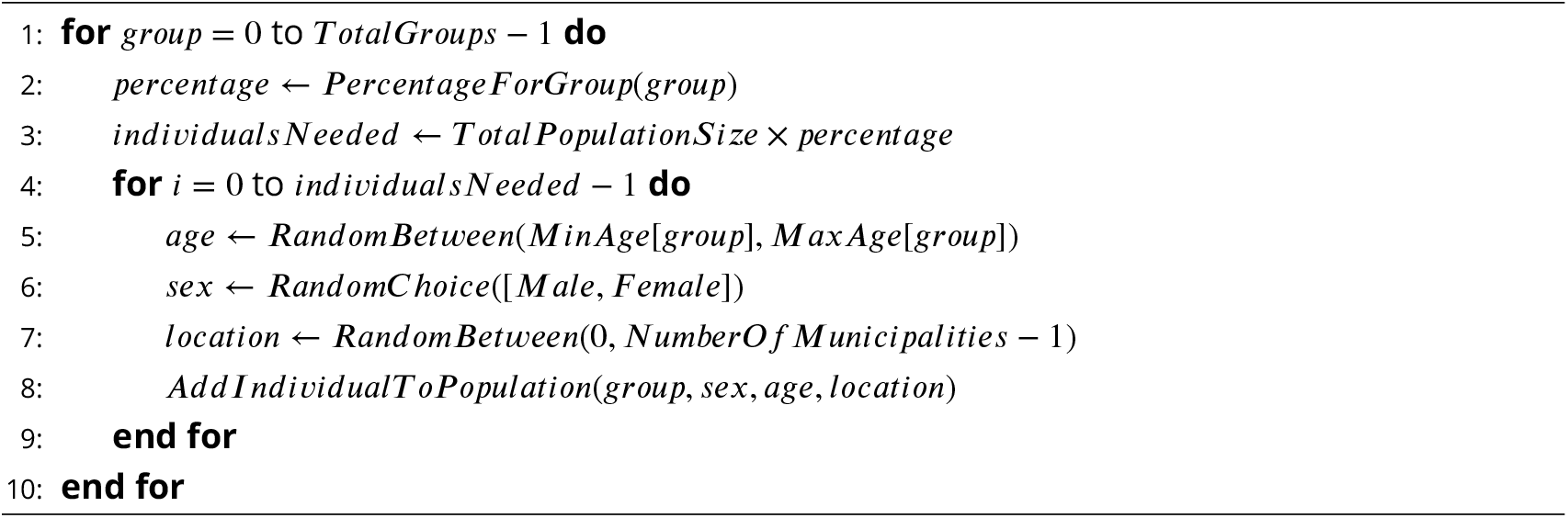

### COVID-19 epidemiological model

In order to study the effects of applying microdata on individual-level agent-based modeling, we use the recent epidemiological case of the COVID-19 spread. We use this case as it is a well-studied recent case for which the agent behaviors are well-defined and scale out to a country-wide population.

The ABM that models the spread of COVID-19 taking into account non-pharmaceutical interventions is based on the work of ***Dekker et al. (2023***). It models the first wave in 2020 in four distinct phases in which the interventions were put in place. Each phase change introduces changes in the mobility and interactions that take place between agents. The two main behaviors that are followed by each agent are 1) the traveling behavior (see Algorithm 2) and 2) the infection behavior (see Algorithm 3). The traveling behavior updates the location of an agent at each timestep, where the location is one of the 380 municipalities in the Netherlands (in 2020). For each agent, an hourly travel schedule is generated. The hourly travel schedules are based on a gravity model ***Ramos (2016***), which differs from the implementation in ***Dekker et al. (2023***). There are two types of traveling behavior, frequent (traveling during weekdays) and incidental (traveling in the weekend). The exact gravity model used to generate a mobility matrix for frequent and incidental travelers is shown in the following set of equations

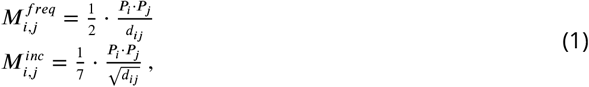

where *d*_*ij*_ is the Euclidean distance between municipality *i* and *j*, which respectively have a population of *P*_*i*_ and *P*_*j*_. The mobility matrix for incidental travelers is adjusted by a factor of 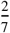 and therefore 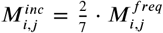. This adjustment reflects the weekends, accounting for two out of the seven days of the week when most people in the Netherlands travel to destinations other than their typical routes (e.g. going out to another city, visiting family or friends, etc.). This differentiation ensures the matrix accurately represents the travel patterns of frequent versus incidental travelers. A weekly schedule is generated per agent on an hourly basis using a Dirichlet distribution which parameters are based on the mixing matrices as explained in ***Dekker et al. (2023***).

The infection behavior follows from a compartmental SEIR model that includes an additional state of ‘hospitalization’ to keep track of agents that are admitted to a hospital as a consequence of a COVID-19 infection. In Algorithm 3, *λ* stands for the force of infection, and *β* is a phase-bound parameter that represents behaviors such as wearing a face mask and social distancing that cannot easily be modeled in an agent-based model. Upon the start of each consecutive phase in the first COVID-19 wave, the interventions that are put into place affect the mobility of the agents, their demographic mixing behavior, and school closure. For more model details, we refer the reader to ***Dekker et al. (2023***).

We initialize the agent with data from Statistics Netherlands (CBS) to be able to mimic demographic mixing as realistically as possible. As an element in its normal operations as the Dutch National Statistical Institute, CBS continuously and automatically updates its databases. Demographic datasets are typically updated with a monthly cadence but other types of data can be collected depending on the specific data collection. From the ‘basis-registratie personen’ (BRP), the base register data on this Dutch population, we use the variables sex, age, work status, education, and residence to categorize agents into demographic groups according to ***Dekker et al. (2023***). The social interaction between agents in this model is determined by a mixing matrix that is based on the survey study of ***Prem et al. (2017***).

Another aspect in which our model differs from the model presented in ***Dekker et al. (2023***) is the increased resolution of which an agent represents an aggregate number of citizens. In contrast to just modeling a 1:100 agent-to-person ratio, we explore a range of ratios up to 1:1 in order to understand the benefits of high-resolution agent-based modeling on the model outcomes, with a special focus on the accuracy of these outcomes.

A list of all relevant model parameters and their values can be found in Appendix 1.

### Calibration

In order to allow for the calibration of large agent-based models in BioDynaMo, we developed a new method that can be used for distributing multiple simulations as separate processes using MPI (Message Passing Interface), the Multi-Simulation Manager. Any iterative algorithm that can run its iterations independently of each other can be used within the simulation manager, but for the scope of this study we focus on the Particle Swarm Optimization (PSO) algorithm ***Kennedy and Eberhart (1995***). The choice for the PSO algorithm was motivated by the fact that our objective function is non-convex and non-differentiable.

#### Algorithm 2

Traveling behavior

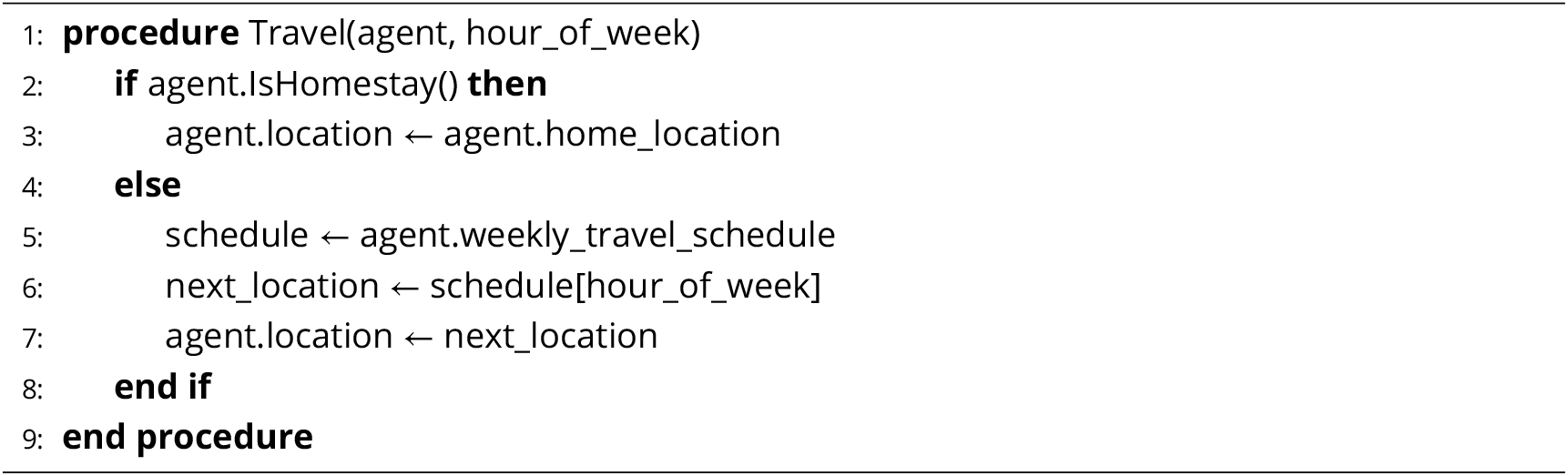

#### Algorithm 3

Infection behavior

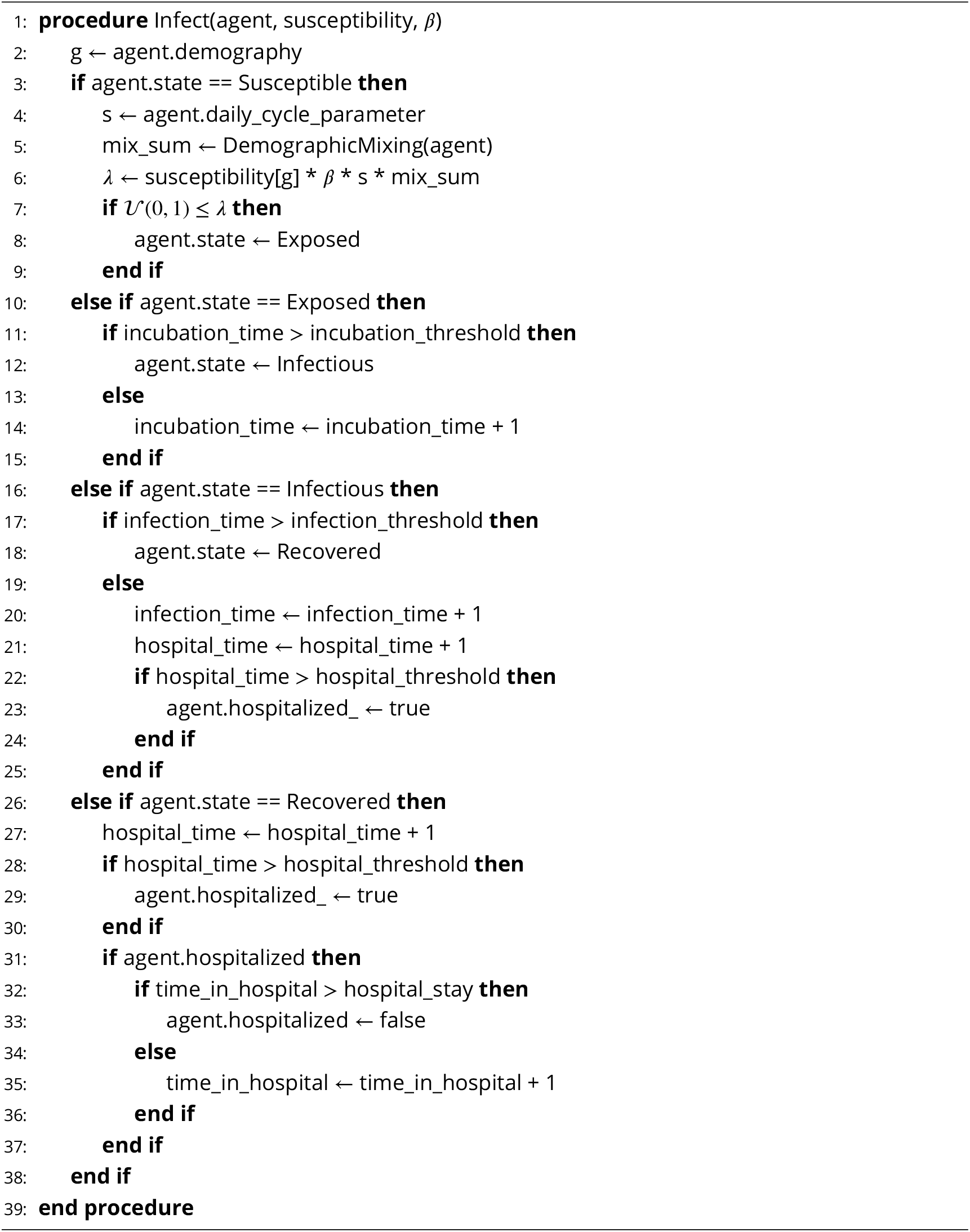

In short, PSO is a population-based optimization algorithm used to find the global optimum solution in a search space. In PSO, a set of particles is initialized randomly within the search space, and each particle moves in the space to find the global optimum. The movement of each particle is guided by its own best-known position and the global best position found so far by the entire swarm. The algorithm continues until the convergence criteria are met or a maximum number of iterations is reached. PSO is a widely used optimization algorithm due to its simplicity and efficiency. Note that here the particles are not the agents of the model: in the present context, each particle represents a separate full-scale simulation with 17.4 million agents (i.e. a resolution of one agent representing one person).

Figure 7 shows an overview of the implementation of the Multi-Simulation Manager. In the context of parallel computing, MPI facilitates the coordination of multiple processors by assigning each a unique identifier, known as a *rank*. The implementation of the Multi-Simulation Manager uses a master-worker paradigm, where one rank (the master rank) oversees the computational work needed to be executed by the worker ranks. In the figure, the master rank initiates a PSO algorithm with *N* particles and at most *max*_*iter* iterations. Each particle, within each iteration, represents a full simulation and is executed by one of the worker ranks in the worker pool, and is repeated *M* times for statistical significance. The simulation is executed in a multi-threaded fashion with OpenMP with a user-specified number of threads. Each worker rank computes the mean-squared error (MSE) between the simulated values and the observed (expected) values. After a full iteration, the master receives the MSE from each worker and updates the particle positions using the optimization algorithm. The algorithm stops when convergence is reached or *max*_*iter* is reached.

**Figure 7.**
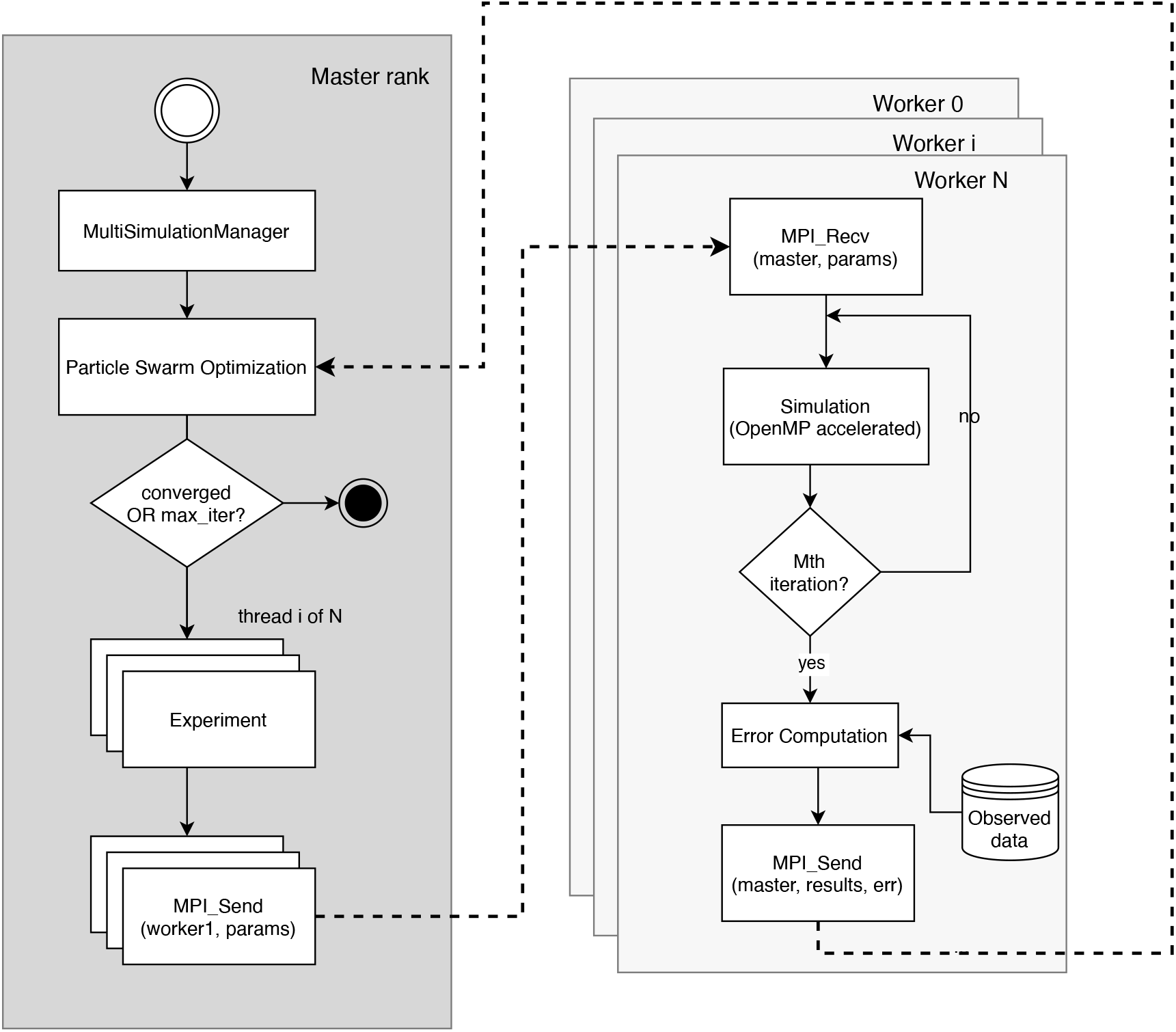
Flowchart of the Multi-Simulation Manager using the Particle Swarm Optimization algorithm for calibrating against observed data

For this case study, we calibrate our agent-based model based on the initial doubling time of observed hospital admissions during the first wave of the COVID-19 pandemic in the Netherlands. This approach aligns with the methods described in the study by Dekker et al. (2022), which highlights the common practice in epidemiology of calibrating models to the doubling phase of a pandemic. Specifically, the doubling time refers to the period during which the number of cases or hospitalizations doubles, providing a critical measure of the virus’s spread rate. In this instance, the doubling time spanned from March 13th to March 27th, approximately two weeks.

The objective of the PSO algorithm, in this case, is minimizing the error of the simulated hospital admissions and the observed hospital admissions during the doubling period, which accounts for approximately 15% of the total observed hospital admission dataset. The consequent course of the hospital admissions after the doubling period is left to be predicted by the model. As such, the model should be able to predict the peak of the wave (both in time and in amplitude) as well as the decline after the peak.

### Parameter Sensitivity Analysis

Understanding the influence of each free parameter on the model’s outcome is crucial for assessing the stability of an agent-based model. Parameters that significantly alter the outcome with only minor adjustments might indicate sensitive dependencies within the model, warranting further investigation to understand their implications fully. This sensitivity could reflect critical dynamics within the model but could also suggest areas requiring more robust definition or validation. Conversely, if a parameter anticipated to have a strong correlation with the model output exhibits little variation during sensitivity analysis, this may indicate that the parameter is not adequately constrained by the available data. Such findings should prompt a reevaluation of both the parameter settings and the data used for model calibration.

There exist various methods to perform parameter sensitivity analysis on agent-based models ***ten Broeke et al. (2016***). In order to choose the right method it is important to know what one is trying to achieve with the results of the analysis. In this work, we wish to quantify the variability of the four beta parameters as described in COVID-19 epidemiological model on the correspondence of the simulated hospital admission to the observed data. The resulting outcome should give us an indication of the robustness of the chosen model parameters and the relative influence on the model outcome among them. The authors of ***ten Broeke et al. (2016***) recommend starting such an analysis with a version of the one-at-a-time (OAT) method. Although the OAT method is meant to uncover the emergent patterns and mechanisms of an ABM, the goal of quantifying the variability of the four beta parameters tells us about the emergence of the pandemic’s peak hospital admission value and its location in time. These values describe the global emergence of the COVID pandemic that our case study revolves around.

#### One-at-a-Time Method

With the one-at-a-time (OAT) or the one-factor-at-a-time (OFAT) method one varies one model parameter while the other model parameters stay the same. The variation occurs typically around a nominal value which is the value that best corresponds to the desired model outcome. The difference between the OAT and OFAT methods is the range of parameter variability: the range in OAT is narrow, while the range in OFAT is wide. The authors of ***ten Broeke et al. (2016***) state that the narrow range examination (OAT) can be used to estimate the partial derivatives of the model outcome with respect to the parameter in question, whereas OFAT aims to show the form of relationship between the outcome and the parameter. The latter is of interest at present because of the need to explore as much as possible the range of outcomes that can be represented. The former is also important for future use of these models since that technique can be used to determine which parameters policymakers or national response teams should try to influence, and in which direction, for the most effective pandemic control.

### Secure Supercomputing Infrastructure

Processing confidential CBS microdata requires a secure computing environment in order to avoid data leakage. The ODISSEI Secure Supercomputer (OSSC) by ***Scheerman et al. (2021***) provides such an environment. The OSSC runs on a virtualized partition of the Dutch national supercomputer, Snellius, and is connected to the CBS data infrastructure using a dedicated VPN connection. CBS curates various person-level datasets which are made available in pseudonomized form to researchers under specific guidelines. The microdata used in this work, as referred to in Table 2, is one such dataset.

In order to allow for our simulations to be reproducible among various computing platforms, we containerized the simulation code and all its dependencies in an Apptainer (formerly known as Singularity) container. This allowed us to prototype and run the model in BioDynaMo on other computing platforms before running full-scale simulations and calibrations on the OSSC. Since the microdata is only available within the OSSC, we used the synthetic population data for our prototyping efforts as described in Synthetic population data.

### Code

This work uses the BioDynaMo agent-based simulation software to model and simulate the COVID-19 model. The model code is available at https://github.com/Senui/covid-abm-paper. Compiling this code requires a custom BioDynaMo build. For reproducibility purposes, we created an Apptainer (formerly known as Singularity) container (***Hesam (2024***)), which can be used to reproduce the results in this paper. The instructions to obtain and run the simulations can be found in Appendix 2.

## Data Availability

All data produced in the present work are contained in the manuscript

## Acknowledgments

The authors extend their gratitude to the Netherlands Organisation for Scientific Research (NWO) for funding the compute resources that facilitated the experiments conducted in this study. We thank SURF (www.surf.nl) for the support in using the National Supercomputer Snellius. We also thank the CBS Microdata Services team for their support and guidance in utilizing these resources.

## Appendix 1 Model details

**Appendix 1—table 1.** Input Parameters for the COVID-19 model.

| Parameter | Value | Description |
| --- | --- | --- |
| repeat | 10 | Number of times to repeat the simulation |
| population_size | 17181084 | Total population size |
| init_infection_rate | 0.1 | Initial infection rate |
| init_infection_time | 408 hours | Initial infection time |
| total_hours | 2256 | Total hours from 27 Feb to 1 June 2020 |
| phase_1_hours | 336 | Hours between 27 Feb and 12 March |
| phase_2_hours | 264 | Hours between 12 March and 23 March |
| phase_3_hours | 1176 | Hours between 23 March and 11 May |
| phase_4_hours | 504 | Hours between 11 May and 1 June |
| phase_2_mobility_reduction | 0.372 | Mobility reduction in phase 2 |
| phase_3_mobility_reduction | 0.424 | Mobility reduction in phase 3 |
| phase_4_mobility_reduction | 0.201 | Mobility reduction in phase 4 |
| beta1 | 0.0701 | Transmission rate parameter for phase 1 |
| beta2 | 0.0093 | Transmission rate parameter for phase 2 |
| beta3 | 0.3533 | Transmission rate parameter for phase 3 |
| beta4 | 0.2380 | Transmission rate parameter for phase 4 |
| phase_2_homeschooling_parents | 12% | Percentage of parents homeschooling in phase 2 |
| no_fixed_seed | false | Whether a fixed seed is used for simulation |
| incubation_shape_param | 20 | Shape parameter for the incubation period |
| incubation_scale_param | 110.4 | Scale parameter for the incubation period (hours) |
| infection_shape_param | 1 | Shape parameter for the infection period |
| infection_scale_param | 120 | Scale parameter for the infection period (hours) |
| hospitalization_shape_param | 14 | Shape parameter for hospitalization period |
| hospitalization_scale_param | 240 | Scale parameter for hospitalization period (hours) |
| hospital_average_mean | 2.48 | Mean of hospital stay (days) |
| hospital_average_sigma | 0.913 | Standard deviation of hospital stay |
| homestay_mean | 15 | Mean duration of staying at home (days) |
| homestay_sigma | 6 | Standard deviation of staying at home duration |

## Appendix 2 Reproducing results

This paragraph describes the steps needed to reproduce the results of this work. Some of the results require access to sensitive datasets provided by Statistics Netherlands as referred to in Table 2, which can only be accessed in a secure remote environment. See the corresponding footnote in Table 2 for more information.

For the purpose of reproducibility, we have created an Apptainer container ***Hesam (2024***) that can be downloaded and used to build and run the simulation code in, which is available at https://github.com/Senui/covid-abm-paper. Apptainer is an open-source software tool that can be installed from https://apptainer.org. We recommend a Linux-based system to perform the simulations on.

Once Apptainer is installed in the simulation environment and the Apptainer image is downloaded, the BioDynaMo source code and the simulation code need to be cloned from the aforementioned Github repository. The simulation environment can then be set up as shown in as follows:

**Listing 1.**
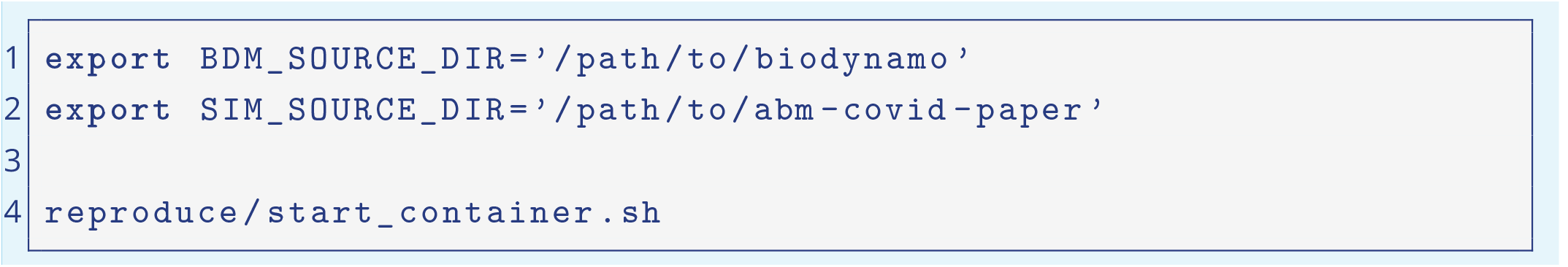
Running a single simulation

This will deploy a containerized environment that is identical to the environment with which the results for this paper were produced. It will automatically start a simulation with the input parameters defined in the bdm.json. To run any other set of input parameters you can adjust the contents of the bdm.json file. There are various configuration files present in the simulation code repository (in the config directory), which are named according to the type of experiments that are run for this paper. The results of a single simulation can be found in the directory build/output/single-experiments

To run multiple simulations distributed on a cluster or supercomputer (for calibration or parameter sensitivity analysis) the following script needs to be invoked:

**Listing 2.**
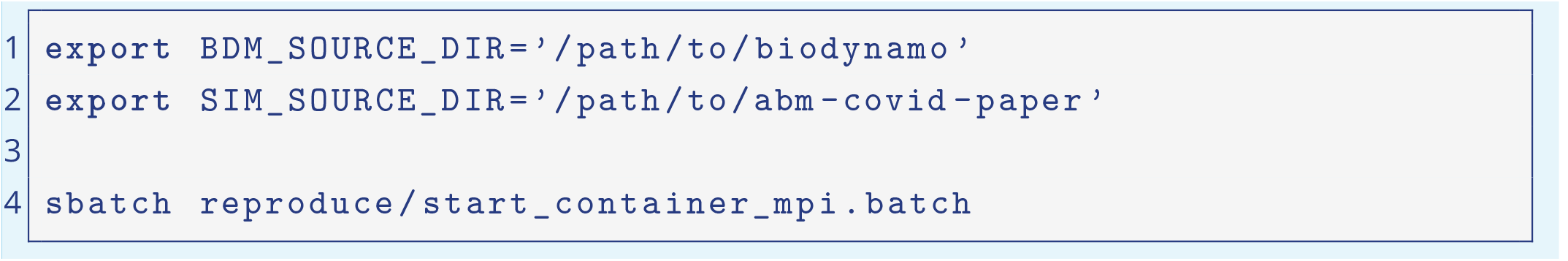
Running many simulations distributed

The results of multiple simulation runs can be found in the directory build/output/experiments_*<*timestamp*>*.

## Footnotes

1 Non-public microdata from Statistics Netherlands. Under certain conditions, these microdata are accessible for statistical and scientific research. For further information.

